# Prevalence of previous infection with SARS-CoV-2 and persistent symptoms at a large university

**DOI:** 10.1101/2021.07.08.21260201

**Authors:** Mark H. Ebell, David Forgacs, Ye Shen, Ted M. Ross, Cassie Hulme, Michelle Bentivegna, Hannah B. Hanley, Alexandria M. Jefferson, Lauren Haines

## Abstract

**Importance:** Universities are unique settings with large populations, congregate housing, and frequent attendance of events in large groups. However, the prevalence of previous infection with SARS-CoV-2 in university students, including symptomatic and asymptomatic disease, is unknown.

**Objective:** To determine the prevalence of previous infection, risk factors for infection, and the prevalence of persistent symptoms following infection among university students.

**Design:** This was a cross-sectional study that surveyed students about demographics, risk factors, and symptoms, and simultaneously tested their saliva for IgA antibodies to SARS-CoV-2. To estimate the prevalence of previous infection we adjusted our intentional sample of a diverse student population for year in school and age to resemble the composition of the entire student body, and adjusted for the imperfect sensitivity and specificity of the antibody test. Univariate and multivariate analysis was used to identify independent risk factors for infection.

**Setting:** A large public university in Athens, Georgia between January 22 and March 22, 2021.

**Participants:** Undergraduate and graduate students; 488 completed the survey, 432 had a valid antibody result. and 428 had both.

**Exposure:** Previous infection with SARS-CoV-2 based on measurement of IgA antibodies in saliva and adjustment for sample characteristics and test accuracy.

**Main Outcomes and Measures:** The primary outcome was the estimated prevalence of previous infection with SARS-CoV-2. Secondary outcomes were independent risk factors for infection, and the prevalence of persistent symptoms among persons reporting a previous symptomatic infection.

**Results:** The estimated prevalence of previous infection for 432 participants with valid antibody results was between 41% and 42%. Independent risk factors for infection included male sex, having a roommate with a known symptomatic infection, and having 2 or fewer roommates. More frequent attendance of parties and bars was a univariate risk factor, but not in the multivariate analysis. Of 122 students reporting a previous symptomatic infection, 14 (11.4%) reported persistent symptoms a median of 132 days later.

**Conclusions and Relevance:** Previous infection with SARS-CoV-2, both symptomatic and asymptomatic, was common at a large university. Measures that could prevent resurgence of the infection when students return to campus include mandatory vaccination policies, mass surveillance testing, and testing of sewage for antigen to SARS-CoV-2.

**Key Points:** *Question:* What is the prevalence of previous infection with SARS-CoV-2 and the prevalence of persistent symptoms in university students?

*Findings:* In this sample of 432 students who provided saliva for IgA antibodies, we estimate that 41% to 42% had evidence of previous infection. Of 122 reporting a previous symptomatic infection, 14 (11%) were still symptomatic a median of 132 days later.

*Meaning:* Symptomatic and asymptomatic infections with SARS-CoV-2 are common among university students, and a significant percentage had persistent symptoms over a long duration.

## Introduction

It has become clear that many patients infected with SARS-CoV-2 are asymptomatic or minimally symptomatic.^1,2^ This is an important factor in the spread of the virus through the human population, as these persons can still spread the disease to others.^3,4^ Therefore, understanding the extent of asymptomatic infection is important to inform future disease control efforts.

Universities are unique settings with large populations, congregate housing, and frequent attendance of events in large groups such as sporting events, classrooms, bars, and other venues. A narrative review of cohort studies found that younger cohorts tended to have higher rates of asymptomatic infection with SARS-CoV-2.^2^ This was confirmed by a large cohort study of 5484 quarantined case contacts, which found that younger persons were significantly more likely to experience an asymptomatic infection than older persons. For example, only 22% of individuals 20 to 39 years of age developed a symptomatic infection compared to 65% of people over the age of 80 years.^5^

As of May 26, 2021 a total of 6,391 symptomatic cases have been reported at the University of Georgia in Athens, Georgia, USA (16.3% of the 39,147 person student body). Of these, 4756 (12.1%) occurred during 2020.^6^ In a previous study, we used data from reported symptomatic cases and surveillance testing of asymptomatic students to estimate that ∼75% of SARS-CoV-2 infections were asymptomatic.^6^ However, this estimate requires confirmation via population-based testing for antibodies to SARS-CoV-2.

Typically, serum samples are used for antibody detection via enzyme-linked immunosorbent assays (ELISAs); however, saliva samples have also been used successfully for the detection of SARS-CoV-2 antibodies.^7,8^ IgG antibody profiles from plasma and saliva are highly correlated for other pathogens as well.^9^ Using IgA antibody-based SARS-CoV-2 serological tests, the majority of antibodies detected in saliva are mucosal^10,11(p20)^ We therefore set out to determine the prevalence of previous symptomatic and asymptomatic infection with SARS-CoV-2 in a sample of students at the University of Georgia using detection of IgA antibodies in saliva. We also sought to identify independent predictors of previous infection, as well as the proportion of students who had persistent symptoms.

## Methods

This was a cross-sectional study. It was approved by the Human Subjects Committee at the University of Georgia (Version 00000889, Project 00003338). All participants gave written informed consent prior to participation. Participants were not compensated for participation; internal funding was used to purchase laboratory supplies.

### Population

The University of Georgia is a large public institution in Athens, Georgia, USA with 29,765 undergraduates and 9,382 graduate and professional students enrolled in the fall semester of 2020. Students were required to be on campus during the fall semester, and faculty were encouraged to teach in person unless they obtained an Americans with Disabilities Act exemption approved by Human Resources at the University. Our goal was to identify a sample of 500 students from all levels that was representative of the student body. Because testing for SARS-CoV-2 was not required at any point during the semester, we instead obtained a list of classes that included the names of instructors, the college, the number of students allowed in the class, and the instructional level. We identified classes with at least 20 students from each of the university’s 17 colleges, selecting both early undergraduate, late undergraduate, and graduate classes to develop as representative a sample as possible. We then asked instructors whether we could come to their class to identify students who would voluntarily provide saliva samples and who would complete a one-page survey. Data collection was done by trained student volunteers.

### Processing of saliva samples

Approximately 1 mL of saliva was collected in Bead Ruptor tubes with 1.4 mm ceramic beads (Omni International, Kennesaw, GA, USA). These tubes were pre-filled with 0.5 mL of viral transport medium containing Hanks balanced salt solution (HBSS) with 2% FBS, 100μg/mL gentamicin, and 0.5μg/mL Amphotericin B, as well as 60μl of 1x cOmplete^™^ Protease Inhibitor Cocktail (Roche, Basel, Switzerland). Following collection, saliva samples were immediately placed in a cooler, and within one hour were refrigerated at 4° C. They were centrifuged at 250 rpm for 30s and stored at −20°C. Immediately before use, the saliva samples were heat inactivated for 45 minutes in a 56°C water bath to destroy any infectious SARS-CoV-2 virus in the sample.^12^

### Enzyme-linked immunosorbent assay (ELISA)

Immulon^®^ 4HBX plates (Thermo Fisher Scientific, Waltham, MA, USA) were coated with 100 ng/well of recombinant SARS-CoV-2 RBD protein in PBS and left overnight at 4°C in a humidified chamber. Plates were blocked using 2% bovine serum albumin (BSA) Fraction V (Thermo Fisher Scientific, Waltham, MA, USA) and 1% gelatin from bovine skin (Sigma-Aldrich, St. Louis, MO, USA) in PBS/0.05% Tween20 (Thermo Fisher Scientific, Waltham, MA, USA) at 37°C for 90 min. Samples were diluted to a final saliva concentration of 1:8 using blocking buffer, and incubated overnight at 4°C in a humidified chamber. Plates were washed 5 times with PBS/0.05% Tween20. IgA antibodies were detected using horseradish peroxidase (HRP)-conjugated goat anti-human IgA detection antibody (Southern Biotech, Birmingham, AL, USA) at a 1:4000 dilution, and incubated for 90 min at 37°C. Plates were then washed 5 times with PBS/0.05% Tween20 prior to development with 100μL of 0.1% 2,2’-azino-bis(3-ethylbenzothiazoline-6-sulphonic acid) (ABTS, Bioworld, Dublin, OH, USA) solution with 0.05% H_2_O_2_ for 18 minutes at 37°C. The reaction was terminated with 50μL of 1% (w/v) SDS (VWR International, Radnor, PA, USA). Colorimetric absorbance was measured at 414nm using a PowerWaveXS plate reader (Biotek, Winooski, VT, USA). All samples were run in duplicate and the mean of the two blank-adjusted optical density (OD) values were used in downstream analyses. IgA equivalent concentrations were calculated based on a 7-point standard curve generated by a human IgA reference protein (Athens Research and Technology, Athens, GA, USA).

### Determination of cutoffs for defining a positive antibody test

The ELISA test for SARS-CoV-2 IgA antibodies was administered simultaneously with a serum test for SARS-CoV-2 IgG antibodies as a reference standard to 18 volunteers. The optimal cutoff that maximizes sensitivity and specificity at 89% was based on Youden’s J statistic. ^13^ The cutoff between positive and negative saliva samples was determined to be > 61.85 ng/mL, and that threshold was used to define a positive or negative test for the univariate and multivariate analysis.

### Survey

The survey was completed during each class period, and included questions about year of study, age, sex, race, ethnicity, type of housing, and number of room or housemates. Students were also asked whether they had experienced a previous confirmed COVID-19 infection, whether they were ever symptomatic, and whether they were still symptomatic. Finally, they were asked about adherence to face coverings and distancing, frequency of attendance of bars and parties, and whether someone they lived with had tested positive. All samples and surveys were identified by a code number only. Surveys were administered from approximately January 22 to March 22 of 2021. Data collection was halted as vaccination began among students to avoid confounding results (students were also asked to not provide a sample if they had been vaccinated).

### Analysis to estimate prevalence

Sample prevalence was weighted by gender and year in school to reflect an appropriate distribution among UGA students (UGA Factbook: https://oir.uga.edu/_resources/files/factbook/pages/UGAFactBook_p20.pdf). The weighted prevalence was then further adjusted using the approach proposed by Diggle,^14^ as the saliva test had imperfect sensitivity. A cutoff of > 68.21 ng/ml was chosen for estimation of prevalence because it had a specificity of 100%, therefore requiring only adjustment for sensitivity rather than for both sensitivity and specificity (see Appendix Table). We also chose a slightly higher cutoff of > 72.82 ng/ml (also 100% specific) as a sensitivity analysis for the estimation of prevalence.

### Analysis for predictors of positive antibody test

A multivariate logistic regression model was used to assess the association between student characteristics such as demographics and behaviors and COVID-19 infection. Because of skewed distributions for some variables and small sample sizes for some categories, categories for race, year in school, and number of people sharing a household were combined for the multivariate analysis. The antibody test result was the dependent variable with age, sex, race, year in school, housing, number of people sharing a household, mask wearing off campus, numbers of parties and bars, and roommates positive as independent variables. Results are presented as adjusted odds ratios and 95% confidence intervals.

### Descriptive analysis of participants with history of confirmed symptomatic COVID-19

A descriptive analysis was done to look at students reporting symptoms for a previous laboratory-confirmed positive viral test for SARS-CoV-2 infection as well as the presence of persistent symptoms at the time of survey. The length of time participants had persistent symptoms was estimated using the reported date of their positive COVID test and the midpoint of our survey data collection, February 22, 2021.

## Results

A total of 497 participants were enrolled between January 20 and March 22, 2021, of whom 8 did not submit a survey, one submitted a survey but no saliva sample, and 62 who provided indeterminate or inadequate saliva samples. This left a total of 488 participants who completed the survey, 432 participants with a valid antibody result. and 428 who provided both (Table 1). The mean age of our study population was 20.6 years, 68.7% were female, and 85.3% identified as white. A previous laboratory-confirmed positive viral test for COVID-19 was reported by 140 of 488 students (28.7%).

**Table 1.**
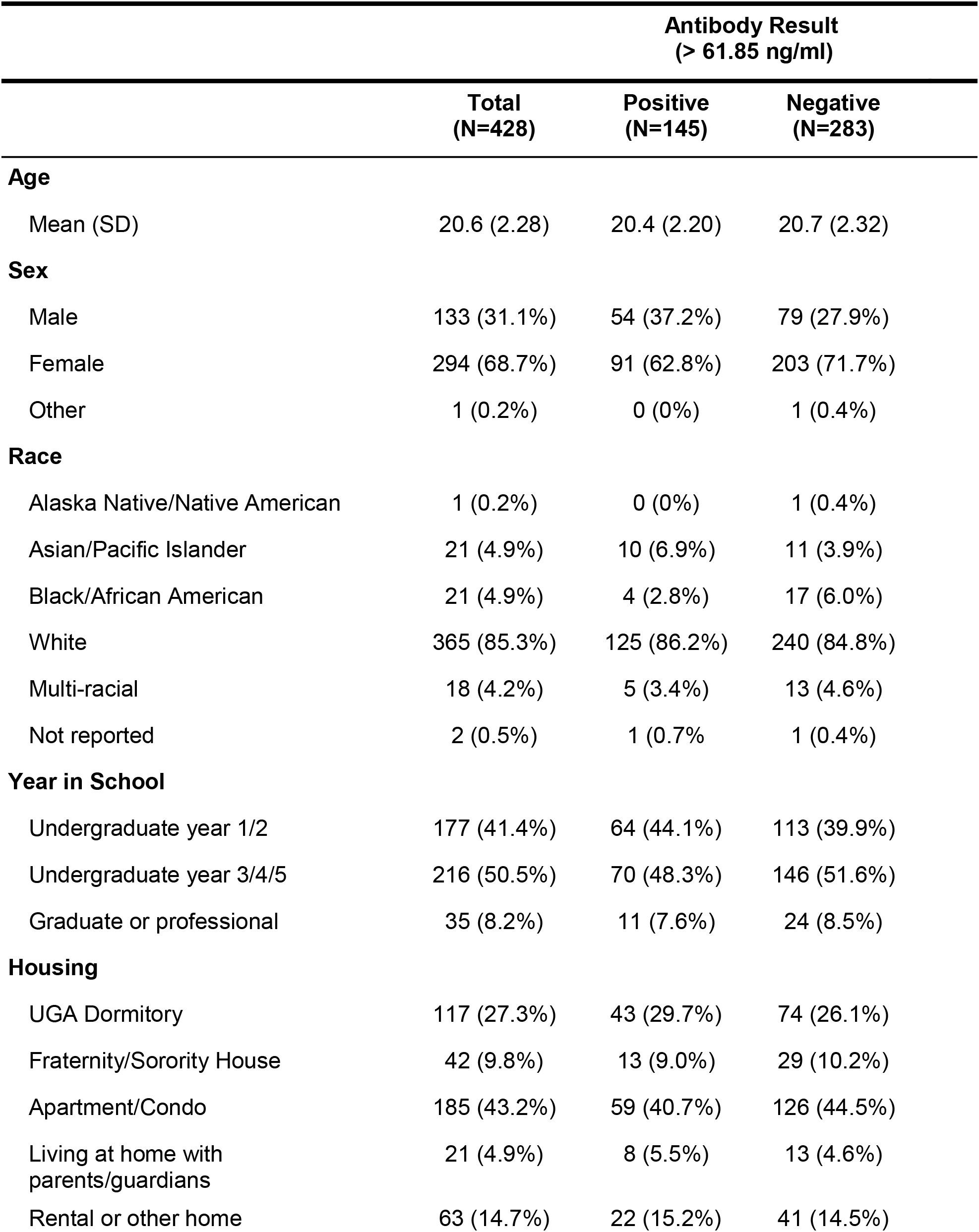
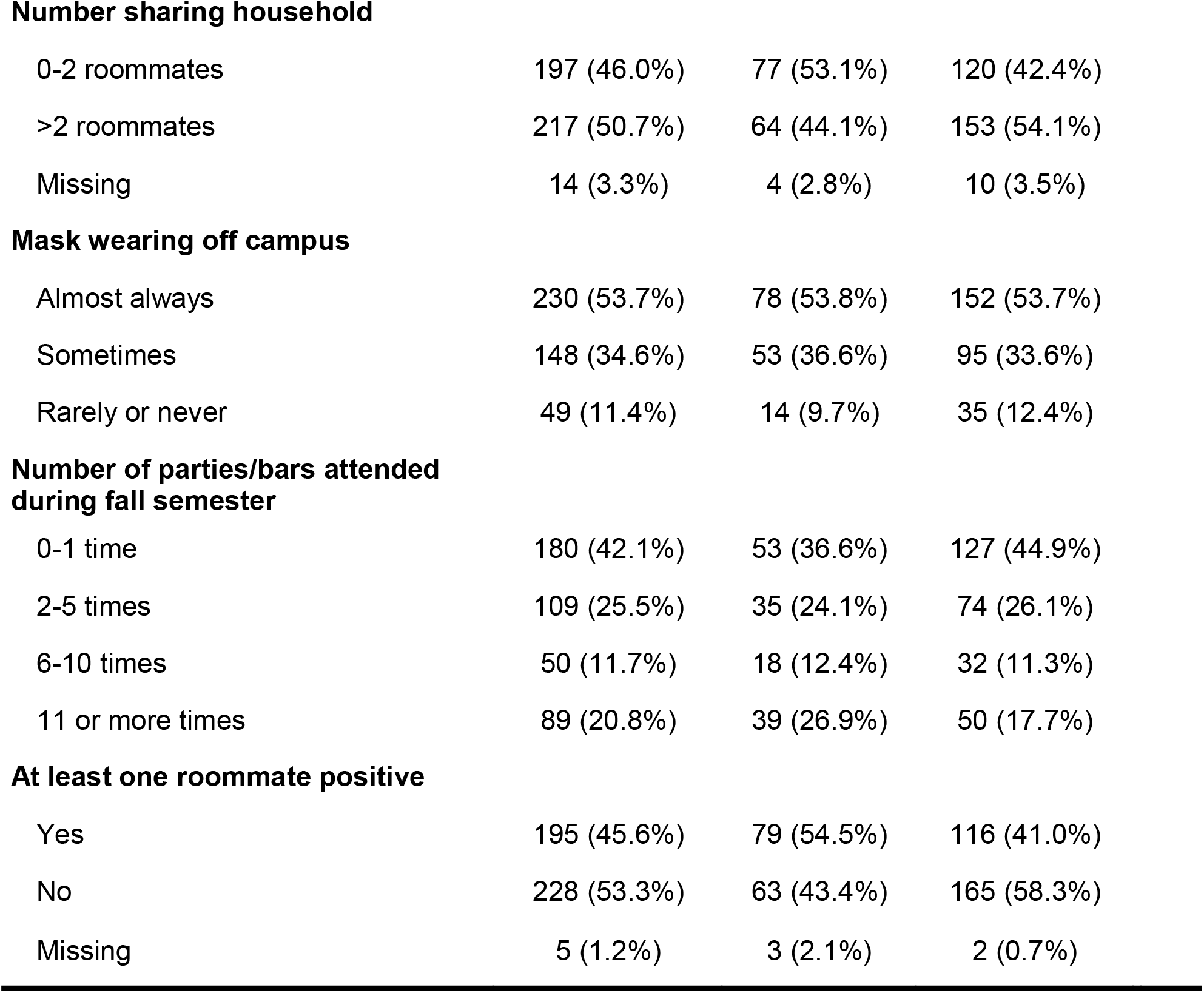
Characteristics of study participants with both valid antibody result and completed survey (n=428)

|  | **Antibody Result**<br>**(> 61.85 ng/ml)** |  |  |
| --- | --- | --- | --- |
|  | **Total**<br>**(N=428)** | **Positive**<br>**(N=145)** | **Negative**<br>**(N=283)** |
| **Age** |  |  |  |
| Mean (SD) | 20.6 (2.28) | 20.4 (2.20) | 20.7 (2.32) |
| **Sex** |  |  |  |
| Male | 133 (31.1%) | 54 (37.2%) | 79 (27.9%) |
| Female | 294 (68.7%) | 91 (62.8%) | 203 (71.7%) |
| Other | 1 (0.2%) | 0 (0%) | 1 (0.4%) |
| **Race** |  |  |  |
| Alaska Native/Native American | 1 (0.2%) | 0 (0%) | 1 (0.4%) |
| Asian/Pacific Islander | 21 (4.9%) | 10 (6.9%) | 11 (3.9%) |
| Black/African American | 21 (4.9%) | 4 (2.8%) | 17 (6.0%) |
| White | 365 (85.3%) | 125 (86.2%) | 240 (84.8%) |
| Multi-racial | 18 (4.2%) | 5 (3.4%) | 13 (4.6%) |
| Not reported | 2 (0.5%) | 1 (0.7%) | 1 (0.4%) |
| **Year in School** |  |  |  |
| Undergraduate year 1/2 | 177 (41.4%) | 64 (44.1%) | 113 (39.9%) |
| Undergraduate year 3/4/5 | 216 (50.5%) | 70 (48.3%) | 146 (51.6%) |
| Graduate or professional | 35 (8.2%) | 11 (7.6%) | 24 (8.5%) |
| **Housing** |  |  |  |
| UGA Dormitory | 117 (27.3%) | 43 (29.7%) | 74 (26.1%) |
| Fraternity/Sorority House | 42 (9.8%) | 13 (9.0%) | 29 (10.2%) |
| Apartment/Condo | 185 (43.2%) | 59 (40.7%) | 126 (44.5%) |
| Living at home with<br>parents/guardians | 21 (4.9%) | 8 (5.5%) | 13 (4.6%) |
| Rental or other home | 63 (14.7%) | 22 (15.2%) | 41 (14.5%) |
| **Number sharing household** |  |  |  |
| 0-2 roommates | 197 (46.0%) | 77 (53.1%) | 120 (42.4%) |
| >2 roommates | 217 (50.7%) | 64 (44.1%) | 153 (54.1%) |
| Missing | 14 (3.3%) | 4 (2.8%) | 10 (3.5%) |
| **Mask wearing off campus** |  |  |  |
| Almost always | 230 (53.7%) | 78 (53.8%) | 152 (53.7%) |
| Sometimes | 148 (34.6%) | 53 (36.6%) | 95 (33.6%) |
| Rarely or never | 49 (11.4%) | 14 (9.7%) | 35 (12.4%) |
| **Number of parties/bars attended during fall semester** |  |  |  |
| 0-1 time | 180 (42.1%) | 53 (36.6%) | 127 (44.9%) |
| 2-5 times | 109 (25.5%) | 35 (24.1%) | 74 (26.1%) |
| 6-10 times | 50 (11.7%) | 18 (12.4%) | 32 (11.3%) |
| 11 or more times | 89 (20.8%) | 39 (26.9%) | 50 (17.7%) |
| **At least one roommate positive** |  |  |  |
| Yes | 195 (45.6%) | 79 (54.5%) | 116 (41.0%) |
| No | 228 (53.3%) | 63 (43.4%) | 165 (58.3%) |
| Missing | 5 (1.2%) | 3 (2.1%) | 2 (0.7%) |

**Table 2.** Results of multivariate logistic regression with presence of antibodies (> 61.85 ng/ml) as the dependent variable.

| **Risk factor** | **Adjusted odds ratio (95% CI)** | **p** |
| --- | --- | --- |
| Age (years) | 0.97 (0.84-1.13) | 0.698 |
| Male sex | 1.66 (1.03 – 2.67) | 0.036 * |
| Non-white race | 1.05 (0.49 – 2.26) | 0.896 |
| Year in school |  |  |
| Undergraduate year 1/2 | REF |  |
| Undergraduate year 3/4/5 | 0.92 (0.46 – 1.82) | 0.803 |
| Graduate | 0.62 (0.18 – 2.17) | 0.453 |
| Housing |  |  |
| Dormitory | REF |  |
| Greek | 1.08 (0.38 – 3.04) | 0.884 |
| Apartment | 1.57 (0.70 – 3.50) | 0.273 |
| At home | 2.68 (0.87 – 8.22) | 0.085 |
| Rental home | 2.24 (0.82 – 6.11) | 0.117 |
| Masks wearing off campus |  |  |
| Almost always | REF |  |
| Sometimes | 1.01 (0.61 – 1.67) | 0.968 |
| Rarely or never | 0.56 (0.26 - -1.20) | 0.134 |
| Number of parties and bars attended during fall semester |  |  |
| 0 to 1 | REF |  |
| 2 to 5 | 1.11 (0.63 – 1.99) | 0.708 |
| 6 to 10 | 1.19 (0.55 – 2.55) | 0.658 |
| 11+ | 1.65 (0.86 – 3.15) | 0.130 |
| At least one roommate positive | 1.92 (1.18 – 3.12) | 0.009 * |
| Number sharing housing > 2 | 0.41 (0.23 – 0.72) | 0.002 * |

\* p < 0.05

### Estimate of overall prevalence

Among all 432 participants who provided saliva samples and had a deterministic measure of IgA titers, the raw prevalence estimate was 30.0% (95%CI: 26.0-34.6%) using the cutoff for prevalence estimation of >68.21 ng/ml. Most of these participants were undergraduate students (n=392), and they had a raw prevalence of 30.4% (95%CI: 26.0-35.1%). After weighting by their gender and year in school, the weighted prevalence was 31.9% (95%CI: 24.4-39.4%). Finally, with adjustment to account for the imperfect sensitivity of the saliva test, the estimated adjusted prevalence was 41.0% (95%CI: 31.4-50.7%). The analysis was repeated using a cutoff of > 72.82 ng/ml as a sensitivity analysis and the adjusted prevalence was estimated to be 42.2% (31.4-52.9%).

### Univariate and multivariate analysis of risk factors for infection

In the univariate analysis, a positive antibody test was significantly more likely in male students (40.6% vs 30.8%, p = 0.048), in students reporting more than 5 attendances at bars or parties during the semester (41.0% vs 30.5%, p = 0.03), in those with at least one roommate who had been diagnosed with COVID-19 (40.5% vs 27.6%, p = 0.01), and in those reporting 2 or fewer people sharing their residence (39.1% vs 29.5%, p = 0.04). There was no significant difference in the likelihood of a positive antibody test based on mask wearing off campus, type of housing, race, or year in school.

The multivariate analysis showed that participants who shared housing with more than two roommates during the fall 2020 semester were less likely to have received a positive COVID antibody test (OR 0.43, 95% CI 0.24, 7.74). Students who lived with roommates that had also received a confirmed COVID diagnosis were more likely to be positive for COVID antibodies (OR 2.01, 95% CI 1.23, 3.32). Social activities that include mask wearing off campus and the number of parties or bars attended did not have a significant association with a positive antibody result.

### Prevalence of persistent symptoms

Of the 488 participants who returned a survey, 140 participants had received a previous laboratory confirmed diagnosis of COVID (Table 3). Additional analysis of these patients showed that 122 (87.1%) of the participants were experiencing symptoms at the time of diagnosis, and 14 (11.4%) of participants with laboratory confirmed symptomatic COVID reported still having symptoms from their infection at the time that the survey was taken. The median duration of persistent symptoms in these 14 patients was 132 days, with a range of 12 to 253 days; 13 of 14 reported symptoms for more than 1 month, and 10 for more than 2 months.

**Table 3.** Characteristics of participants with previous confirmed COVID-19 infection

|  | Confirmed Covid<br>(N=140) |
| --- | --- |
| **Symptoms** |  |
| Yes | 122 (87.1%) |
| No | 18 (12.9%) |
| **Persistent Symptoms** |  |
| Yes | 14 (10.0%) |
| No | 126 (90.0%) |
| **Number of Days with Persistent Symptoms (n=14)** |  |
| Mean (SD) | 132 (22.5) |
| Median [Min, Max] | 131 [12, 253] |

### Comparison of previous viral testing and antibody results

In the 428 students with both survey and antibody results, 85 of 112 (75.9%) reporting a previous laboratory confirmed positive viral test also were positive for salivary IgA antibodies. Of 316 students without a previous laboratory confirmed positive viral test, 60 (18.9%) were positive for salivary IgA antibodies.

## Discussion

We estimate that 41% of students had experienced a previous infection with SARS-CoV-2 at our university. Since it takes approximately 2 weeks for IgA to become detectable in saliva,^15^ the previous infections detected by our study represent infections from the beginning of the pandemic until approximately early February. Based on published data there were 5160 symptomatic cases reported during that period,^16^ whereas we estimate that the total number of cases was approximately 16,050 (95% CI 12,292 to 19,847) in the student population of 39,147 persons. Therefore, over two-thirds of infections in university students were asymptomatic.

The extent of asymptomatic transmission has important policy and public health implications. Measures like temperature checks and apps that ask about symptoms are very unlikely to have an impact when so many infected and infectious persons at a university are asymptomatic. Measures like mandatory vaccination for students, faculty and staff,^17^ testing of sewage for antigen to SARS-CoV-2,^18^ and regular testing of the entire population^19^ are more likely to be effective. While many students have antibodies due to previous infection, it is not clear how durable that immunity is. Also, vaccination rates are low among young persons. For that reason, the American College Health Association recently recommended requiring COVID-19 vaccination for all students on campus in the fall of 2021.^20^

Independent predictors of infection include male sex, having a roommate who had a confirmed symptomatic infection, and having fewer than two roommates. While the number of parties and bars attended during fall semester was linearly and significantly associated with the likelihood of infection, it was not an independent predictor in the multivariate analysis. It is not clear why having fewer roommates increased the risk of previous infection. While we speculated that perhaps those with more roommates had larger “pods” and therefore felt less need to go to parties and bars, that was not the case (percentage attending more than 5 parties or bars in the semester was 34% for those with 2 or fewer roommates and 30% for those with 2 or more, p = 0.43). There may also be residual confounding with dormitory as a place of residence, which accounted for more than half of those reporting 2 or fewer roommates.

It is concerning that 11% of students who reported a previous symptomatic infection also reported persistent symptoms a median of 131 days after their initial diagnosis. Based on the total symptomatic reported infections (n=6391), that suggests that approximately 700 students may be experiencing persistent symptoms. This requires further study to assess the longitudinal course of illness, the type of symptoms, and their severity in these “long-haul” patients which has not been previously reported in a population of young adults. A study in healthcare workers found that the prevalence of symptoms declined slowly over time in those with persistent symptoms, but that 15% were still experiencing symptoms 8 months later.^21^

A limitation of our study is the fact that this was not a random sample of students. However, we did deliberately sample a broad range of classes, levels, and colleges to generate a diverse sample, and we also adjusted our estimate to the year and sex distribution of the university as a whole. While the saliva test is less accurate than serology, it was more practical for rapid data collection in a classroom setting, and our analysis was able to adjust for the lack of sensitivity using a validated approach. Assessment of risk factors was by self-report and there may be a degree of social desirability bias to some of the questions, such as adherence to mask-wearing and attending parties or bars. Finally, we did not assess the type or severity of persistent symptoms.

### Conclusions

We estimate that over 40% of students at our university were infected with SARS-CoV-2, and that many were asymptomatic. Risk factors for infection include having an infected roommate and male sex, and approximately 11% with a symptomatic infection were experiencing persistent symptoms months later. Given low vaccination rates among young people, and the increasing prevalence of more contagious variants, mandating vaccination prior to return to campus is needed to prevent resurgence of the infection and harm to students, staff and faculty, especially as many universities have abandoned other protective measures.

## Data Availability

Data are not available as further analysis is ongoing by the co-authors as part of a thesis. They will be made available upon reasonable request when this analysis is completed.

## Acknowledgements

The authors would like to thank Katie Mailloux, Jasmine Burris, Omar Hamwy, and Jordan Byrne for technical assistance. The recombinant proteins were produced by Jeffrey Ecker, Spencer Pierce, Ethan Cooper, and the team in the Center for Vaccines and Immunology protein production core. The authors would also like to thank Emma Calhoun Ellis, Caroline Johnston, Mary Anne E. Roach, Caroline Lindsey, Ansley Connelly, and Yazan Bouchi for their assistance with data collection

