## Appendix for "Prevalence of previous infection with SARS-CoV-2 and persistent symptoms at a large university"

**Appendix. Determination of cutoffs for abnormal tests and for prevalence estimates.**

Appendix Table 1. Sensitivity and specificity of different antibody titer cutoffs for the diagnosis of previous infection with SARS-CoV-2

| **Antibody titer** | **Sensitivity (%)** | **95% CI** | **Specificity (%)** | **95% CI** |
| --- | --- | --- | --- | --- |
| > 22.90 | 100 | 70.09% to 100.0% | 11.11 | 0.5699% to 43.50% |
| > 39.16 | 100 | 70.09% to 100.0% | 22.22 | 3.948% to 54.74% |
| > 43.22 | 100 | 70.09% to 100.0% | 33.33 | 12.06% to 64.58% |
| > 44.62 | 100 | 70.09% to 100.0% | 44.44 | 18.88% to 73.33% |
| > 46.34 | 88.89 | 56.50% to 99.43% | 44.44 | 18.88% to 73.33% |
| > 49.06 | 88.89 | 56.50% to 99.43% | 55.56 | 26.67% to 81.12% |
| > 53.95 | 88.89 | 56.50% to 99.43% | 66.67 | 35.42% to 87.94% |
| > 58.57 | 88.89 | 56.50% to 99.43% | 77.78 | 45.26% to 96.05% |
| **> 61.85** | **88.89** | **56.50% to 99.43%** | **88.89** | **56.50% to 99.43%** |
| > 64.45 | 77.78 | 45.26% to 96.05% | 88.89 | 56.50% to 99.43% |
| **> 68.21** | **77.78** | **45.26% to 96.05%** | **100** | **70.09% to 100.0%** |
| **> 72.82** | **66.67** | **35.42% to 87.94%** | **100** | **70.09% to 100.0%** |
| > 77.32 | 55.56 | 26.67% to 81.12% | 100 | 70.09% to 100.0% |
| > 95.59 | 44.44 | 18.88% to 73.33% | 100 | 70.09% to 100.0% |
| > 140.6 | 33.33 | 12.06% to 64.58% | 100 | 70.09% to 100.0% |
| > 220.4 | 22.22 | 3.948% to 54.74% | 100 | 70.09% to 100.0% |
| > 319.4 | 11.11 | 0.5699% to 43.50% | 100 | 70.09% to 100.0% |

Appendix Figure 1. ROC curve and dot plot for saliva assay.

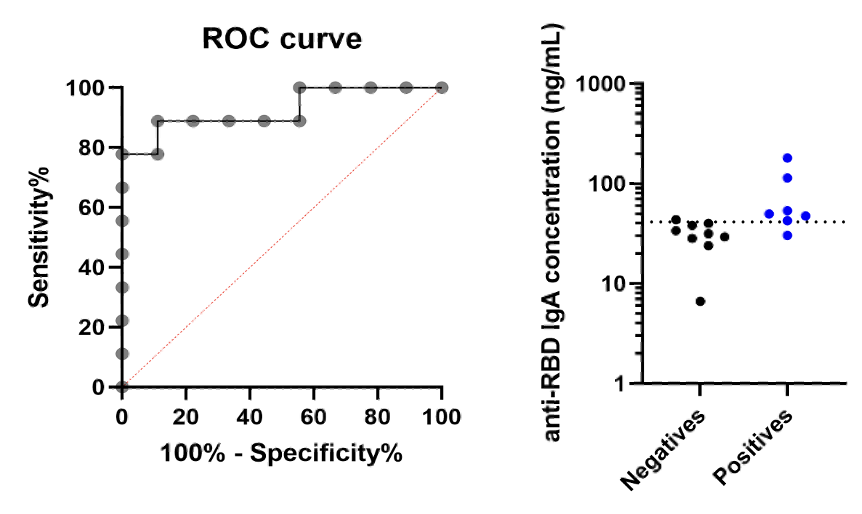
